# Effect of sustained viral clearance on liver-related mortality among individuals living with hepatitis C by treatment era: a population-based retrospective cohort study

**DOI:** 10.1101/2022.10.29.22281693

**Authors:** Aysegul Erman, Karl Everett, William W. L. Wong, Farinaz Forouzannia, Christina Greenaway, Naveed Janjua, Jeffrey C. Kwong, Beate Sander

## Abstract

**Background:** Chronic infection with hepatitis C virus (HCV) is a leading cause of liver-related mortality. Direct-acting antivirals (DAAs) have revolutionised treatment by offering profound improvements in sustained viral clearance (SVR) and tolerability resulting in rapid expansion of treatment for individuals for whom HCV treatment had previously been less feasible, such as those with advanced liver disease or with drug and/or alcohol-related substance use. Given these clinical policy shifts, the primary objective of this study was to assess the impact of SVR on liver-related death among important clinical groups and the secondary objective was to explore changes in predictors of liver-related death by treatment era using real-world data from a large population-based cohort.

**Methods:** We conducted a population-based, linked cohort study of all Ontario residents with HCV viremia between January 1st, 1999, and December 31st, 2018, with follow up to 31st May 2021 (N=73,411). Population-level health administrative, clinical, and demographic data were accessed at ICES. Cause-specific hazard models were used to explore the impact of SVR on liver-related death and to identify factors associated with the rate of liver-related death in the DAA and pre-DAA treatment eras. The moderating effects of liver disease severity and substance-use disorder on the relationship between SVR and liver-related-mortality was explored by stratification.

**Results:** Among Ontario residents diagnosed with living with HCV, the achievement of SVR was associated with a significant reduction in liver related mortality (adjusted hazard ratio [aHR] 0.22, 95%CI: 0.20-0.24 vs. no SVR). This was also observed across progressive liver disease severity levels (aHR 0.13, 95%CI: 0.10-0.17 for individuals without cirrhosis; aHR 0.11, 95%CI: 0.06-0.17 for those with compensated cirrhosis, and aHR 0.24, 95%CI: 0.22-0.27 for those with advanced liver disease vs. no SVR) and by substance use status (aHR 0.24, 95%CI: 0.21-0.27 for those with a history of substance use disorder; and aHR 0.21, 95%CI: 0.18-0.24 for those without vs. no SVR). Additionally, factors such as age at diagnosis, sex, liver disease severity, immigration status, birth year, substance use, HBV-coinfection, viral genotype, and markers of social marginalisation were independent predictors of liver-related mortality. However, sex, and viral genotype no longer displayed significant associations with liver-related death in the DAA era as was observed in the earlier treatment era.

**Conclusions:** This study provides real-world evidence showing profound impact of SVR on liver-related mortality in a population-based sample of individuals with CHC and highlights the importance of early diagnosis and treatment. This study further demonstrates significant mortality benefits of SVR regardless of substance use status highlighting the importance of supporting marginalised individuals in treatment access.

## Introduction

Chronic hepatitis C (CHC) is associated with major morbidity and mortality. Over time, the chronic persistence of hepatitis C virus (HCV) can cause variable degrees of hepatic fibrosis and eventually lead to cirrhosis, and end-stage liver disease necessitating a liver transplant [1–4].

Although the achievement of sustained viral clearance (SVR) has been shown to substantially reduce mortality for individuals living with hepatitis C, the standard-of-care consisted of interferon-based therapy for decades, offering only limited efficacy over a long-treatment duration [5,6]. Moreover, traditional therapies were of limited use for individuals with a history of substance use and mental illness due to their low tolerability and neuropsychiatric side effects [7]. Similarly, tolerability was also a concern for individuals with advanced liver disease [8]. In fact, interferon-based regiments were contraindicated for individuals with decompensated cirrhosis and demonstrated only limited efficacy in treating recurrent infection among immunosuppressed liver-transplant recipients [8,9].

Since the introduction of highly effective direct-acting antivirals (DAAs) in 2013, clinical policy guidelines for CHC have been rapidly changing. Given the significant advantages of DAAs over the traditional interferon-based therapies, individuals for whom earlier therapies had been contraindicated, including those with advanced liver disease or significant comorbidities such as substance use, can now be effectively treated [10–13]. Moreover, compared to earlier therapies, DAAs demonstrate much higher efficacy among post-transplant individuals, making treatment much more feasible [14,15]. In addition, due to the pan-genotypic effectiveness of DAAs, HCV genotyping no longer plays a role in treatment decisions as it did in the pre-DAA era [16]. Finally, the high rates of SVR exhibited by DAAs also offer an opportunity to eliminate HCV as a public health concern and meet the WHO target of a 65% reduction in mortality by 2030 [17]. However, despite rapid shifts in clinical practice, real-world impact of SVR on liver-related death in the DAA era, specifically among important clinical populations such as those with advanced liver disease and substance use is lacking.

Our primary objective was to assess the impact of SVR on liver-related death in the DAA and pre-DAA treatment eras among important clinical population and our secondary objective was to explore changes in predictors of liver-related deaths by treatment era using real-world data from a population-based cohort of individuals living with hepatitis C in the province of Ontario, Canada’s largest jurisdiction.

## Methods

### Study population and setting

In Ontario, Canada, confirmation, and testing of reportable diseases including hepatitis C virus (HCV) and hepatitis B virus (HBV) typically occurs at Public Health Ontario (PHO) laboratories. In the current study, we performed a retrospective cohort study of Ontario residents undergoing confirmatory testing for HCV viremia at PHO laboratories between January 1st, 1999, and December 31st, 2018 (N=79,899). After the exclusion of individuals whose laboratory testing records could not be linked to demographic data of all Ontario residents held at ICES (N=6,488), our study cohort included 73,411 unique individuals with confirmed HCV viremia. We followed the study cohort up to May 31st, 2021, to identify total and liver-related mortality.

### Treatment era

The study cohort consisting of individuals testing positive for HCV RNA in Ontario between January 1999 and December 31st, 2018, were further classified by treatment era into 1) individuals receiving a positive HCV RNA diagnosis during the pre-DAA era (Jan 1999-Dec 2013) and followed up until the end of the pre-DAA era (N=54,854); and 2) individuals receiving a positive HCV RNA diagnosis during the DAA era (Jan 2014-Dec 2018) and followed up until May 31st, 2021 (N=18,557).

### Data sources

We linked health administrative, clinical, and demographic data held at ICES datasets using unique identifiers and analysed the data at ICES. All data related to HBV and HCV laboratory testing were collected from PHO laboratory dataset. Data on HCV antiviral dispensation were sourced from Ontario Drug Benefit (ODB) using drug identification numbers (DIN) listed in S.Table 3 in the appendix. For those with a record of HCV antiviral drug dispensation from public drug plans, SVR was defined as a negative HCV RNA test result at least 12 weeks after the dispensation of the last HCV antiviral drug; those without a record of HCV antiviral drug dispensation from public sources but who had a final negative HCV RNA test result that took place after a positive HCV RNA were presumed to have achieved SVR through access to treatment via private insurance schemes external to ODB. The Ontario Marginalisation Index Database (ONMARG) was used to access information on social marginalisation such as neighborhood residential instability quintile, material deprivation quintile and ethnic concentration quintile. We used Immigration, Refugees and Citizenship Canada (IRCC) permanent Resident Database, which holds data individuals who have been granted permanent resident status in Canada since 1985 to determine immigration status. Demographic data including birth year, gender, age, rurality, and neighborhood income quintile were accessed using the Registered Persons Database (RPD). Clinical information on the history of alcohol and/or drug-related substance use disorder, prior diagnosis of cirrhosis, decompensated cirrhosis (DC), hepatocellular carcinoma (HCC), liver transplant, HIV status, cause of death and comorbidities (Aggregated Diagnosis Groups [ADG] comorbidity classification scheme) were collected from the National Ambulatory Care Reporting System (NACRS), Ontario Health Insurance Program (OHIP), Canadian Institute for Health Information Discharge Abstract Database (DAD), Ontario Mental Health Reporting System (OMHRS), Ontario Cancer Registry (OCR) and Office of the Registrar General Death registry (ORGD) datasets, using the relevant diagnostic and procedural codes listed in S.Tables 1-2 and death codes listed in S.Table 4 in the appendix. For comorbidities, we used the Johns Hopkins Adjusted Clinical Group® (ACG) system to determine the number ADGs using a one-year look-back period preceding HCV RNA diagnosis [18]. Baseline characteristics of the study cohort were assembled at the at time of HCV RNA diagnosis.

### Statistical Analysis

To identify factors associated with liver-related mortality, we used cause-specific hazard models to estimate covariate-adjusted hazard ratios (aHR) accounting for non-liver related death as a competing risk [8]. For the cause-specific hazard models, individuals were followed from index date (first data of HCV RNA positivity on record) to either liver-related death, or until other death or censoring on (December 31st, 2013, for pre-DAA era and on May 31st, 2021, for the DAA-era cohorts) with the competing event treated as censored [9].

Stratified analysis was conducted to account for clinical policy changes with respect to the expanded treatment of individuals with advanced liver disease and drug and/or alcohol-related substance use over the two treatment eras, and to explore the moderating effects of liver disease severity and substance-use on the relationship between SVR and liver-related mortality. The analysis was stratified by the level of liver disease severity at time of HCV diagnosis into 1) no cirrhosis, 2) compensated cirrhosis, and 2) advanced liver disease (HCC, DC or liver transplant) and by the presence or absence of alcohol and/or drug-related substance-use disorder on record.

Incidence of liver-related mortality was estimated as the number of observed clinical events divided by the person-years time at risk of event during the observation period for those with and without SVR. Incidence rates were also estimated for all clinical subgroups.

The hazard models adjusted for potential confounders including age at diagnosis, sex, rurality, birth year, comorbidities, neighborhood income, immigrant status, residential instability level, material deprivation level, ethnic concentration level, HIV or HBV coinfection, HCV genotype, alcohol and drug-related substance use disorder, history of liver transplant, liver disease severity and SVR achievement.

For statistical models, we used complete case analysis by excluding observations with missing data. All statistical analysis were performed with SAS Enterprise Guide 7.15 (SAS Institute, Inc.) using PROC PHREG for hazard models and PROC GENMOD to estimate incidence rates.

## Results

### Characteristics of the study cohort

The study cohort consisted of 73,411 individuals with a positive HCV RNA diagnosis in the province of Ontario who were followed up over a median follow-up time of 10 years (IQR: 6-15 years) (**Table 1**). On average, individuals were 45 years of age at the time of HCV RNA diagnosis, 65% were male, 12% were immigrants, 1.5% were coinfected with HIV, 0.6% were coinfected with HBV and 50% had alcohol and/or drug related substance use disorder during the study time frame. More than half of the study cohort had markers of social marginalisation such as lower income levels, higher levels of residential instability and material deprivation. With respect to liver disease severity, the majority (79%) had no evidence of cirrhosis, 7% had compensated cirrhosis, 10% had DC and 5% had HCC at the time of diagnosis. Individuals were most frequently infected with viral genotype-1 (64%), followed by genotype-3 (21%) and genotype-2 (11%). In total 47% of the cohort received treatment of those 80.5% achieved SVR.

**Table 1.** Demographic characteristics of study cohort stratified by treatment era. Demographic characteristics of the study cohort at the time of RNA diagnosis and characteristics of individuals diagnosed during the pre-DAA era (Jan 1999- Dec 2013) and DAA era (Jan 2014- Dec 2018). Frequencies are calculated after exclusion of missing values. *HBV diagnosis is based on hepatitis b surface antigen (HBsAg) reactivity. *Abbreviations: AB: antibody; RNA: ribonucleic acid; HCV: hepatitis c virus; DAA: direct-acting antiviral; ADG: aggregated diagnostic groups; HBV: hepatitis b virus; DC: decompensated cirrhosis; HCC: hepatocellular carcinoma, HIV: human immunodeficiency virus; HR: hazard ratio; CI: confidence interval; q: quintile; N: number of observations;* SVR: sustained viral clearance.

|  | Study Cohort<br>N=73,411 | TREATMENT ERA |  |
| --- | --- | --- | --- |
|  |  | Pre-DAA<br>[1999-2013]<br>N=54,854 | DAA<br>[2014-2018]<br>N=18,557 |
| **Age in years, mean (SD)** |  |  |  |
| Age at diagnosis | 44.7 (12.9) | 45.0 (12.2) | 43.8 (14.8) |
| **Birth cohort, N (%)** |  |  |  |
| <1945 | 5,513 (7.5) | 4,841 (8.8) | 672 (3.6) |
| 1945-1965 | 41,860 (57.0) | 34,204 (62.4) | 7,656 (41.3) |
| >1965 | 26,038 (35.5) | 15,809 (28.8) | 10,229 (55.1) |
| **Male sex, N (%)** | 47,756 (65.1) | 35,781 (65.3) | 11,975 (64.6) |
| **Rural, N (%)** | 7,468 (10.3) | 5,010 (9.3) | 2,458 (13.4) |
| **Neighborhood income quintile, N (%)** |  |  |  |
| Low (quintiles 1-2) | 41,523 (57.7) | 30,362 (56.4) | 11,161 (61.3) |
| Medium (quintile 3) | 12,510 (17.4) | 9,497 (17.7) | 3,013 (16.5) |
| High (quintiles 4-5) | 17,969 (24.9) | 13,930 (25.9) | 4,039 (22.2) |
| **Residential instability quintile, N (%)** |  |  |  |
| Low (quintiles 1-2) | 16,904 (24.0) | 13,169 (25.0) | 3,735 (21.2) |
| Medium (quintile 3) | 11,240 (16.0) | 8,487 (16.1) | 2,753 (15.7) |
| High (quintiles 4-5) | 42,150 (60.0) | 31,059 (58.9) | 11,091 (63.1) |
| **Material deprivation quintile, N (%)** |  |  |  |
| Low (quintiles 1-2) | 17,680 (25.2) | 13,548 (25.7) | 4,132 (23.5) |
| Medium (quintile 3) | 12,064 (17.2) | 9,161 (17.4) | 2,903 (16.5) |
| High (quintiles 4-5) | 40,550 (57.6) | 30,006 (56.9) | 10,544 (60.0) |
| **Ethnic concentration quintile, N (%)** |  |  |  |
| Low (quintiles 1-2) | 25,205 (35.9) | 17,864 (33.9) | 7,341 (41.8) |
| Medium (quintile 3) | 12,942 (18.4) | 9,461 (17.9) | 3,481 (19.8) |
| High (quintiles 4-5) | 32,147 (45.7) | 25,390 (48.2) | 6,757 (38.4) |
| **Immigrant N (%)** | 8,843 (12.0) | 7,210 (13.1) | 1,633 (8.8) |
| **Substance use disorder (ever), N (%)** | 36,378 (49.6) | 25,221 (46.0) | 11,157 (60.1) |
| **HIV positivity, N (%)** | 900 (1.2) | 760 (1.4) | 140 (0.8) |
| **HBV antigen positivity\*, N (%)** | 463 (0.6) | 358 (0.7) | 105 (0.6) |
| **Aggregated diagnosis group categories, N (%)** |  |  |  |
| 0-3 ADGs | 26,106 (35.6) | 19,157 (35) | 16,004 (86.2) |
| 4-7 ADGs | 29,345 (40.0) | 22,351 (40.8) | 1,009 (5.4) |
| 8-10 ADGs | 11,462 (15.6) | 8,643 (15.8) | 1,028 (5.5) |
| >11 ADGs | 6,410 (8.8) | 4,648 (8.4) | 516 (2.9) |
| **Liver disease severity at diagnosis, N (%)** |  |  |  |
| Non-cirrhotic (NC) | 57,692 (78.6) | 41,688 (76.0) | 13,136 (86.5) |
| Compensated cirrhosis (CC) | 5,091 (6.9) | 4,082 (7.4) | 839 (5.5) |
| Decompensated cirrhosis (DC) | 7,376 (10) | 6,348 (11.6) | 816 (5.4) |
| Hepatocellular carcinoma (HCC) | 3,252 (4.5) | 2,736 (5.0) | 393 (2.6) |
| **Liver transplant, N (%)** | 994 (1.4) | 782 (1.4) | 212 (1.1) |
| **HCV genotype, N (%)** |  |  |  |
| Genotype 1 | 43,268 (64.2) | 32,780 (65.1) | 10,488 (61.4) |
| Genotype 2 | 7,391 (11.0) | 5,958 (11.8) | 1,433 (8.4) |
| Genotype 3 | 13,850 (20.5) | 9,576 (19.0) | 4,274 (25.0) |
| Genotype 4 | 1,384 (2.1) | 1,132 (2.2) | 252 (1.5) |
| Other/mixed | 1,534 (2.3) | 900 (1.8) | 634 (3.7) |
| **Treated, N (%)** | 34,932 (47.6) | 19,047 (34.7) | 7,571 (40.8) |
| **SVR, N (% of treated)** | 28,110 (80.5) | 14,732 (77.3) | 6,269 (82.8) |

### Characteristics of the study cohort by treatment era

The majority (75%) of the study cohort were diagnosed in the pre-DAA era (**Table 1**) The median follow-up time was 7 years (IQR: 4-11 years) for individuals diagnosed and followed up within the pre-DAA-era, and 5 years (IQR: 3-6 years) for those diagnosed in the DAA era. On average, individuals diagnosed in the DAA-era were younger; most were born after 1965 (55% vs. 29% in the pre-DAA era). DAA-era cases also had higher levels of substance use disorder (60% vs. 45%), were less likely to be immigrants (9% vs 13%) and more likely to reside in rural areas (13% vs. 9%). Individuals identified during the DAA-era tended to have milder liver disease (87% vs. 76%) and were less likely to be coinfected with HIV (0.8% vs. 1.4%). Additionally, DAA-era cases were more likely to have acquired viral genotype-3 infections (25% vs 19%). Lastly, SVR was also more frequent among those diagnosed in the DAA-era compared to the earlier treatment era (82% vs. 77%).

### Characteristics of the study cohort by liver disease severity

The study cohort consisted of 57,567 individuals without any record of cirrhosis, 5,058 individuals with compensated cirrhosis, and 10,785 with advanced liver disease (DC and/or HCC) at the time of HCV diagnosis (**S.Table5**). When compared to individuals presenting with mild liver disease those with more advanced liver disease tended to be older at diagnosis (52 vs. 43 years), were more likely to be male (72% vs. 68%), and less likely to have substance use disorder (49% vs. 45%). Finally, SVR was 81% for individuals without cirrhosis, 83% for those with compensated cirrhosis, and 75% for those with more advanced liver disease.

### Characteristics of the study cohort by liver disease severity and treatment era

With respect to treatment era, 25% of individuals with mild liver disease, 20% of individuals with compensated cirrhosis, and 15% of individuals with advanced liver disease were diagnosed in the DAA-era (**S.Table5**). For all levels of liver disease severity, individuals diagnosed in the DAA-era were younger, more likely to reside in lower income neighbourhoods and more likely to have a history of substance use but were less likely to be immigrant. In general, DAA-era improvement in treatment initiation were more pronounced for those compensated cirrhosis (50% vs. 41%) and for those with advanced liver disease (44% vs. 35%). There were also improvements in SVR in the DAA-era among individuals presenting with compensated cirrhosis (96% vs. 77%), and with advanced liver disease (89% vs. 59%) compared the pre-DAA era.

### Characteristics of the study cohort by substance use

The study cohort consisted of 37,033 individuals without and 36,378 individuals with a history of drug and/or alcohol-related substance use disorder (**S.Table6**). Compared to individuals without substance use, those with substance use tended to be younger at diagnosis (41 vs.48 years), were more likely to be male (68% vs. 62%), more likely to be HIV-coinfected (1.6% vs. 0.9%) and tended to have neighbourhood-level markers of social marginalisation including lower income (63% vs. 52%), higher residential instability (68% vs. 52%) and greater material deprivation (63% vs. 52%). Individuals with substance use disorder were also much less likely to be immigrants (3% vs. 21%) but were more likely to present with DC at diagnosis (12% vs. 8%) and were more frequently infected with viral genotype-3 (24% vs. 18%). Treatment rate were also lower among those with substance use disorder (43% vs. 52%), as were SVR levels (73% vs. 87%).

### Characteristics of the study cohort by substance and treatment era

In terms of treatment era, 31% of individuals with substance use disorder and 20% of those without were diagnosed in the DAA era (**S.Table6**). Individuals with substance use diagnosed during the DAA-era were slightly younger than those diagnosed in the pre-DAA era (39 vs. 42 years). Compared to the pre-DAA era, there were also improvements in SVR but only among those without substance use disorder (96% vs. 81%).

### Incidence of liver-related death in the study cohort

Incidence of liver-related death in the study cohort by SVR status and age at diagnosis is illustrated in **Figure 1** for clinical populations assessed. Overall, the incidence of liver-related death in the study population was 4 per 1,000 person-years (PY) [5.9/1,000 PY among those without SVR and 1.5/1,000 PY among those with SVR] (**S.Table7**). The incidence of liver-related death increased noticeably with progressive liver disease from 1.1/1,000 PY among those without cirrhosis [1.7/1,000 PY without and 0.2/1,000 PY with SVR] to 22/1,000 PY among those presenting with advanced liver disease [34.7/1,000 PY without and 9/1,000 PY with SVR]. On average, incidence of liver-death was 4.8/1,000 PY for individuals with a history of drug and/or alcohol related substance use [6.6/1,000 PY without and 1.8/1,000 PY with SVR] and 3.3/1,000 PY among those without substance use [5.3/1,000 PY without and 1.2/1,000 PY with SVR].

**Figure 1.**
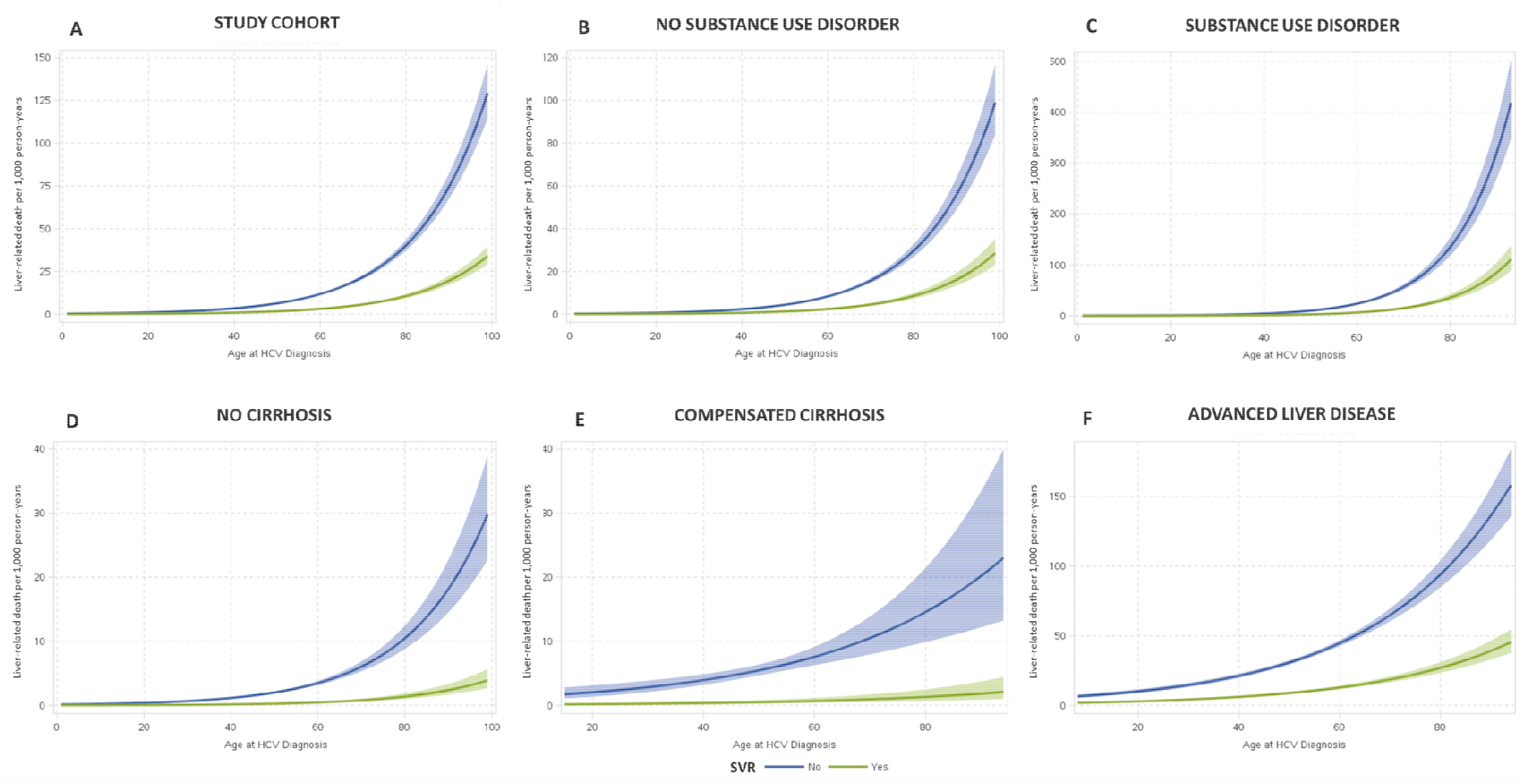
Incidence of liver-related death in individuals diagnosed with hepatitis C by SVR status and age at diagnosis. Figure displays the incidence rate of liver-related death per 1,000 person years by SVR status and age at diagnosis for (A) the total study cohort; (B) those without drug and/or alcohol related substance use disorder; (C) those with drug and/or alcohol related substance use disorder; and by liver disease severity for (D) those without cirrhosis; (E) those with compensated cirrhosis; and (F) those with advanced liver disease. Abbreviations: HCV: hepatitis c virus; SVR: sustained viral clearance.

### Predictors of liver-related death

The predictors of liver-related mortality for the study cohort are displayed in **Table 2**. After adjustment for covariates in the cause-specific hazard model, achievement of SVR was associated with a significant reduction in the rate of liver related mortality (aHR 0.22, 95%CI: 0.20-0.24 vs. no SVR). Additionally, covariates associated with the timing of liver-related death included age (aHR 1.06, 95%CI: 1.06-1.07), liver disease severity at the time of diagnosis (aHR, 2.18, 95%CI: 1.85-2.58 for compensated cirrhosis, aHR 6.34; 95%CI: 6.67-7.08 for DC and, aHR 26; 95%CI 23.9-28.9 for HCC vs. no cirrhosis), sex (aHR 0.75, 95%C,I 0.69-0.82 for female vs male), immigration status (aHR 0.88; 95%CI: 0.78-0.99 for immigrants vs. long-term resident), birth year (aHR 0.58, 95%CI: 0.51-0.6 for those born before 1945, and aHR 0.37, 95%CI: 0.29-0.48 for those born after 1965 vs. individuals born between 1945 and 1965), presence of substance use disorder (aHR 1.56, 95%CI: 1.44-1.68), and viral genotype (aHR 0.84, 95%CI: 0.74-0.95 for viral genotype-2 and aHR 0.78, 95%CI: 0.62-0.97 for viral genotype-4 vs genotype-1).

**Table 2.** Predictors of time-to-liver-related mortality. Predictors of time-to liver-related death following HCV RNA diagnosis. Observations with missing covariates were excluded. Adjusted hazard ratios (HR) were determined using multivariable cause-specific hazard models. Bold values indicate statistical significance at p<0.05. *Abbreviations: SVR: sustained virologic response; AB: antibody; RNA: ribonucleic acid; HCV: hepatitis c virus; DAA: direct-acting antiviral; ADG: aggregated diagnostic groups; HBsAg: hepatitis b surface antigen; HIV: Human immunodeficiency virus; HR: hazard ratio; CI: confidence interval; q: quintile; N: number of observations;* SVR: sustained viral clearance

|  | Study Cohort<br>N=69,558 | TREATMENT ERA |  |
| --- | --- | --- | --- |
|  |  | PRE-DAA<br>N=51,987 | DAA<br>N=17,563 |
| Covariates | Adjusted<br>HR (95% CI) | Adjusted<br>HR (95% CI) | Adjusted<br>HR (95% CI) |
| **Age (yrs.)** |  |  |  |
| At time of diagnosis | **1.06 (1.06-1.07)** | **1.09 (1.09-1.10)** | **1.02 (1.00-1.04)** |
| **Birth year** |  |  |  |
| 1945-1965 [ref.] | ☐ | ☐ | ☐ |
| <1945 | **0.58 (0.51-0.66)** | **0.32 (0.27-0.37)** | 0.85 (0.54-1.32) |
| >1965 | **0.37 (0.29-0.48)** | **0.28 (0.14-0.55)** | **0.22 (0.12-0.40)** |
| **Sex** |  |  |  |
| Male [ref.] | ☐ | ☐ | ☐ |
| Female | **0.75 (0.69-0.82)** | **0.73 (0.67-0.80)** | 0.89 (0.67-1.18) |
| **Rurality** |  |  |  |
| Urban [ref.] | ☐ | ☐ | ☐ |
| Rural | 0.96 (0.84-1.09) | 1.03 (0.89-1.18) | 0.78 (0.51-1.18) |
| **Neighborhood income quintile** |  |  |  |
| Low (q. 1-2) [ref.] | ☐ | ☐ | ☐ |
| Medium (q. 3) | 0.95 (0.85-1.06) | 0.91 (0.81-1.02) | 1.12 (0.77-1.63) |
| High (q. 4-5) | 0.89 (0.79-1.00) | 0.88 (0.78-1.00) | 1.09 (0.77-1.53) |
| **Residential instability quintile** |  |  |  |
| Low (q. 1-2) [ref.] | ☐ | ☐ | ☐ |
| Medium (q. 3) | 0.95 (0.85-1.07) | 0.93 (0.83-1.05) | 1.30 (0.84-2.01) |
| High (q. 4-5) | 0.97 (0.88-1.07) | 0.95 (0.86-1.05) | 1.03 (0.68-1.55) |
| **Material deprivation quintile** |  |  |  |
| Low (q. 1-2) [ref.] | ☐ | ☐ | ☐ |
| Medium (q. 3) | 0.95 (0.85-1.07) | 0.97 (0.86-1.09) | 1.42 (0.98-2.05) |
| High (q. 4-5) | 1.03 (0.92-1.16) | 1.08 (0.96-1.22) | **1.61 (1.11-2.34)** |
| **Ethnic concentration quintile** |  |  |  |
| Low (q. 1-2) [ref.] | ☐ | ☐ | ☐ |
| Medium (q. 3) | 0.99 (0.89-1.10) | 0.97 (0.87-1.08) | 1.16 (0.85-1.60) |
| High (q. 4-5) | **0.91 (0.83-0.99)** | **0.90 (0.82-0.99)** | 1.01 (0.76-1.33) |
| **Immigrant status** |  |  |  |
| Long-term resident [ref.] | ☐ | ☐ | ☐ |
| Immigrant | **0.88 (0.78-0.99)** | 1.04 (0.92-1.19) | 0.87 (0.56-1.35) |
| **Substance use disorder** |  |  |  |
| No [ref.] | ☐ | ☐ | ☐ |
| Yes | **1.56 (1.44-1.68)** | **1.69 (1.55-1.84)** | **1.31 (1.01-1.69)** |
| **HIV positivity** |  |  |  |
| No [ref.] | ☐ | ☐ | ☐ |
| Yes | 0.90 (0.61-1.34) | 0.84 (0.56-1.26) | 0.97 (0.13-6.98) |
| **HBsAG positivity** |  |  |  |
| No [ref.] | ☐ | ☐ | ☐ |
| Yes | **1.57 (1.00-2.47)** | 1.36 (0.83-2.22) | 2.40 (0.75-7.68) |
| **Total number of ADGs (N.)** | **1.01 (1.00-1.02)** | 1.00 (0.99-1.01) | 1.01 (0.97-1.05) |
| **Liver disease at the time of diagnosis** |  |  |  |
| No cirrhosis [ref.] | ☐ | ☐ | ☐ |
| Compensated cirrhosis | **2.18 (1.85-2.58)** | **1.95 (1.64-2.31)** | 1.55 (0.70-3.44) |
| Decompensated cirrhosis | **6.34 (5.67-7.08)** | **4.56 (4.06-5.11)** | **9.15 (5.98-14.0)** |
| Hepatocellular carcinoma | **26.2 (23.9-28.9)** | **16.8 (15.2-18.5)** | **83.5 (59.5-117)** |
| **Liver transplant** |  |  |  |
| No transplant [ref.] | ☐ | ☐ | ☐ |
| Transplant | **1.76 (1.53-2.04)** | **1.68 (1.43-1.97)** | **2.21 (1.58-3.09)** |
| **Viral genotype** |  |  |  |
| Genotype-1 [ref.] | ☐ | ☐ | ☐ |
| Genotype-2 | **0.84 (0.74-0.95)** | **0.83 (0.73-0.95)** | 1.13 (0.78-1.64) |
| Genotype-3 | 1.07 (0.97-1.18) | **1.22 (1.10-1.36)** | 0.82 (0.60-1.10) |
| Genotype-4 | **0.78 (0.62-0.97)** | **0.75 (0.59-0.95)** | 1.06 (0.50-2.27) |
| Other/mixed | 0.94 (0.73-1.20) | 0.81 (0.61-1.09) | 0.76 (0.47-1.24) |
| **Achievement of SVR** |  |  |  |
| No [ref.] | ☐ | ☐ | ☐ |
| Yes | **0.22 (0.20-0.24)** | **0.28 (0.25-0.31)** | **0.26 (0.19-0.35)** |
| **NUMBER OF EVENTS** | **3,116** | **2,836** | **330** |

Moreover, there were important differences with respect to predictors of liver-related death by treatment era. Upon stratification and follow up by treatment era, several factors including sex, viral genotype, ethnic concentration, and the presence of compensated cirrhosis at diagnosis no longer displayed an association with liver-related death in the DAA era as was observed in the earlier treatment era. However, material deprivation was identified as a predictor (aHR 1.61 95%CI: 1.11-2.34) in the DAA-era.

### Predictors of liver-related death by liver disease severity

In the stratified analysis, the achievement of SVR was again associated with a significant lower rate of liver-related death across all levels of liver disease severity for individuals presenting with no cirrhosis (aHR 0.13, 95%CI: 0.10-0.17), those with compensated cirrhosis (aHR 0.11, 95%CI: 0.06-0.17) and those presenting with advanced liver disease at the time of HCV diagnosis (aHR 0.24, 95%CI: 0.22-0.27) **(Table 3)**. Covariates consistently associated with liver-related death across all stages of liver disease included age at diagnosis, birth year, sex, presence of drug and/or alcohol related substance use disorder, comorbidities, and achievement of SVR. Higher income was associated with a lower rate of liver death only for those presenting with no cirrhosis at diagnosis (aHR 0.59 95%CI: 0.451-0.78 vs. low level). Immigrant status was associated with a lower rate of liver death (aHR 0.86 95%CI: 0.75-0.99 vs. long-term resident), and HBV coinfection was associated with a higher liver death (aHR 1.65 95%CI: 1.01-2.72) for those presenting with advanced liver disease.

**Table 3.**
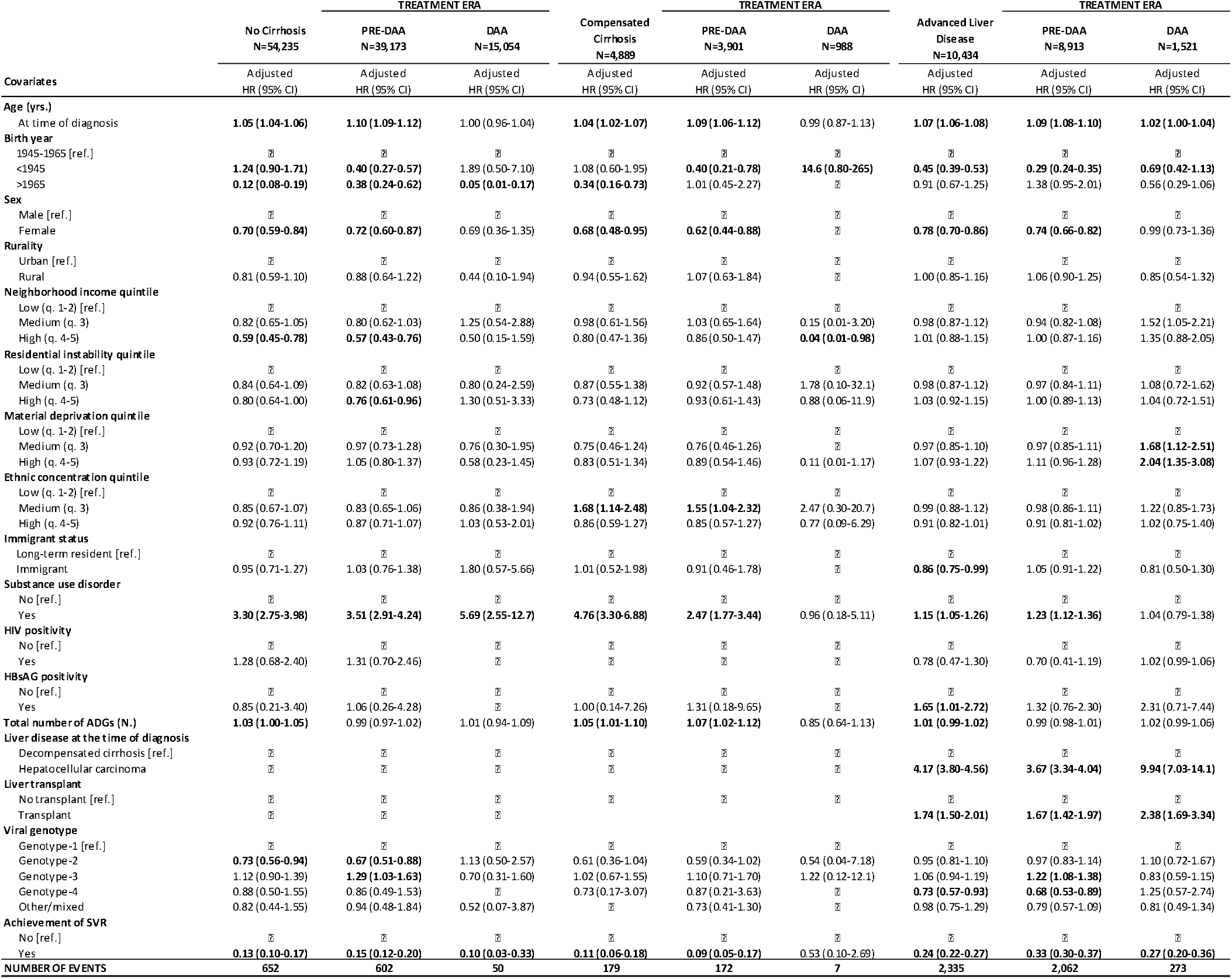
Predictors of time-to-liver-related mortality stratified by liver disease severity. Predictors of time-to liver-related death following HCV RNA diagnosis by liver disease severity. Observations with missing covariates were excluded. Adjusted hazard ratios (HR) were determined using multivariable cause-specific hazard models. Bold values indicate statistical significance at p<0.05. *Abbreviations: SVR: sustained virologic response; AB: antibody; RNA: ribonucleic acid; HCV: hepatitis c virus; DAA: direct-acting antiviral; ADG: aggregated diagnostic groups; HBsAg: hepatitis b surface antigen; HIV: Human immunodeficiency virus; HR: hazard ratio; CI: confidence interval; q: quintile; N: number of observations;* SVR: sustained viral clearance.

Additionally, there were several notable differences in terms of predictors of liver-related death by liver disease levels across treatment eras. In the DAA-era, the presence of alcohol and/or drug related substance use was no longer a predictor of liver-related mortality for individuals with more advanced liver disease while material deprivation was found to be a predictor of liver death among individuals presenting with advanced liver disease. Although, the DAA-era analysis of liver-related death among the non-cirrhotic and more specifically compensated cirrhosis groups are likely to be underpowered given low event rates.

### Predictors of liver-related death by substance use

The effect of SVR on liver-related mortality was similar regardless of individual’s substance use history resulting in significantly lower rates of liver-related death for individuals with substance use (aHR 0.24, 95%CI: 0.21-0.27) as well as for individuals without (aHR 0.21, 95%CI: 0.18-0.24) **(Table 4)**. The covariates associated with liver-related death for those with and without substance use included age at diagnosis, birth year, sex, liver disease severity at diagnosis, genotype, and achievement of SVR. However, HBV coinfection was a predictor of liver-related death for individuals with substance use (aHR 2.76, 95%CI: 1.65-4.59).

**Table 4.**
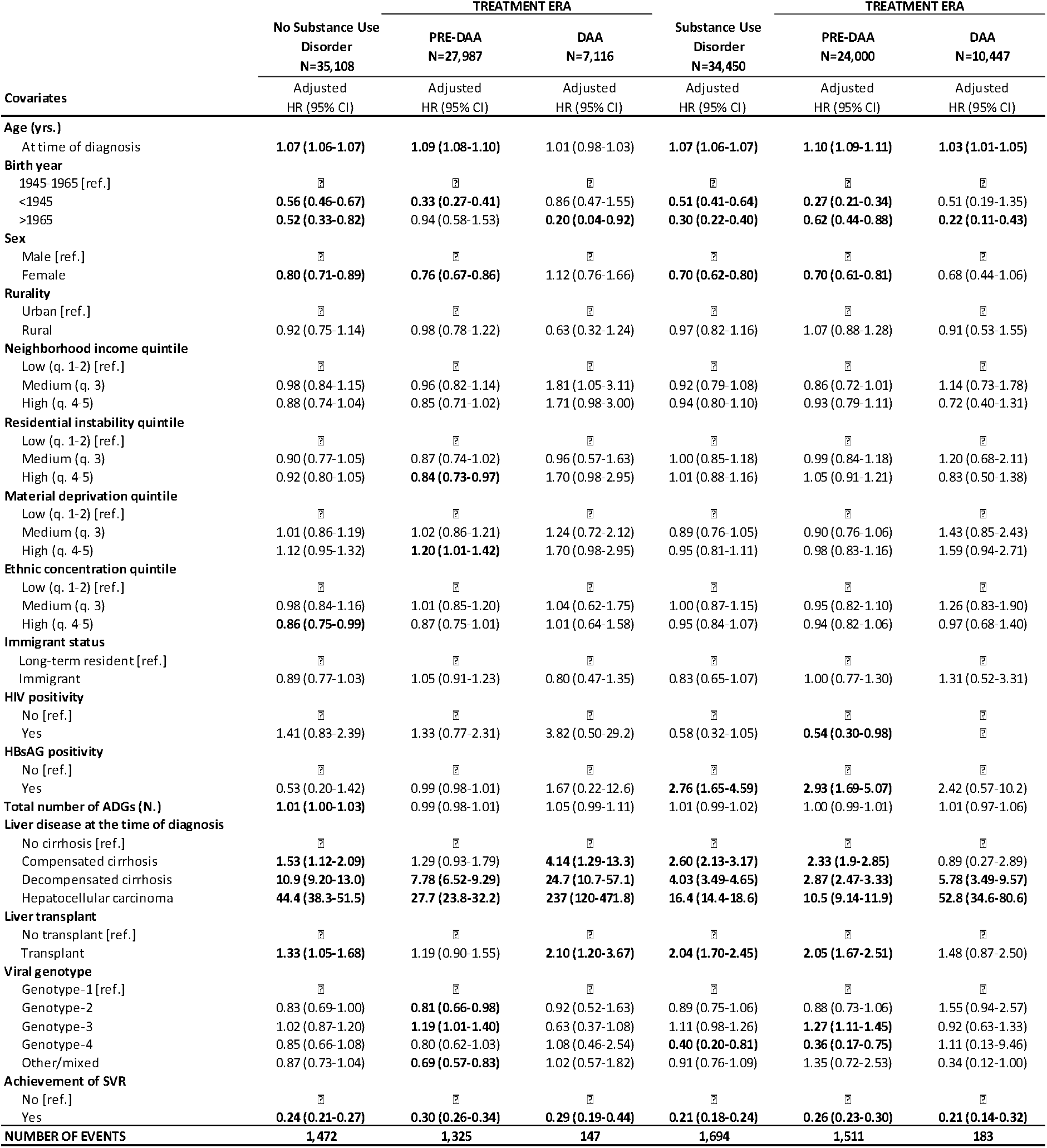
Predictors of time-to-liver-related mortality stratified by substance use. Predictors of time-to liver-related death following HCV RNA diagnosis for those with or without alcohol and/or drug related substance use disorder. Observations with missing covariates were excluded from the analysis. Adjusted hazard ratios (HR) were determined using multivariable and univariate cause- specific hazard models respectively. Bold values indicate statistical significance at p<0.05. *Abbreviations: SVR: sustained virologic response; AB: antibody; RNA: ribonucleic acid; HCV: hepatitis c virus; DAA: direct-acting antiviral; ADG: aggregated diagnostic groups; HBsAg: hepatitis b surface antigen; HIV: Human immunodeficiency virus; HR: hazard ratio; CI: confidence interval; q: quintile; N: number of observations.;* SVR: sustained viral clearance.

There were also differences in predictors by treatment eras. For instance, in the DAA era, sex and viral genotype were no longer a significant predictor of liver death for either group. Similarly, residential instability and material deprivation were no longer predictors of liver death for individuals without substance use, and HBV-coinfection was no longer a predictor of liver death for those with substance use in the DAA era.

## Discussion

There have been major shifts in clinical policy guidelines for CHC following the introduction of DAAs resulting in expanded treatment access to individuals for whom HCV treatment had previously been less feasible. Despite rapid changes, real-world effects of SVR and other important predictors of liver-related death has not been assessed. The current study investigates real-world predictors of liver-related death in the DAA and pre-DAA treatment eras among individuals living with hepatitis C in a population-based cohort in Canada.

We found SVR to significantly reduce liver-related death independent of other covariates and across all clinical populations assessed by liver disease severity and the presence of drug and/or alcohol-related substance use. These findings were largely congruent with a previous U.S study by Backus et al whereby achievement of SVR was independently associated with lower rate of all-cause death among those with advanced liver disease (defined by FIB-4 score over 3.25) receiving DAA treatment (HR 0.26, 95%CI: 0.22-0.31 vs. non SVR) [21]; and a pre-DAA era multicenter study by Van der Meer et al among individual with advanced hepatic fibrosis or cirrhosis (HR 0.26, 95%CI: 0.14-0.49 vs. non SVR) [22]. Moreover, our findings were also consistent with more generalizable population-level real-world studies, including a DAA-era UK study which found that achievement of SVR among individuals with compensated cirrhosis to be associated with lower rates of liver-related death (HR 0.13; 95%CI: 0.05-0.34 vs. non SVR) and all-cause mortality (HR: 0.30; 95%CI: 0.12-0.76 vs. non SVR)[23]. Finally, a recent Canadian study from British Colombia also found SVR to be associated with significant reduction in liver-related deaths (HR 0.22, 95%CI: 0.18–0.27) as well as with all-cause (HR 0.19; 95%CI: 0.17–0.21) and drug-related mortality (HR 0.26, 95%CI: 0.21–0.32) in broad population-based cohort [24]. However, the literature investigating the effects of SVR on mortality specifically among those with alcohol or drug-related substance use is limited. To our knowledge, our study is the first to evaluate the mortality effects of SVR among individuals with drug and/or alcohol related substance use disorder using real world population-based cohort study and to provide evidence that individuals with substance use disorder can realise significant mortality benefits with respect to liver-related death following achievement of SVR.

Moreover, we identified factors such as age at diagnosis, sex, liver disease severity at diagnosis, immigration status, birth year, substance use, HBV-coinfection viral genotype and markers of social marginalisation as independent predictors of liver-related mortality in addition to achievement of SVR. However, we also found important differences by treatment era, whereby factors such as sex, viral genotype, no longer displayed significant associations with liver-related death in the DAA era as was observed in the earlier treatment era. Given the profound mortality impact of liver disease severity, improvements in early diagnosis prior to the development of advanced liver disease and timely access to highly effective therapies will be needed to reduce liver-related mortality in the province.

Our study has some limitations. First, individuals with drug and/or alcohol related substance use disorder are defined as those with a clinical diagnosis of substance use, thus individuals with either less severe or undisclosed substance use are likely to be misclassified. Second, the definition of SVR was based on a record of HCV antiviral dispensation based on public drug plan data. To account for individuals who may have received treatment outside of the provincial drug plan those without a record of HCV antiviral drug dispensation from public sources but who have a final negative test result were assumed to have achieved SVR; thus, a small number of individuals with acute resolved HCV infection may be misclassified as having achieved SVR related viral clearance. Third, because PHO HCV testing data covers test performed between 1999 to 2018, the analyses lack data on those diagnosed prior to 1999 and likely misclassifies individuals who may have achieved SVR after 2018. Moreover, follow-up period of individuals identified in the DAA-era is shorter than those diagnosed in the pre-DAA era.

However, our study has several strengths. We investigated real-world predictors of liver-related death in the DAA and pre-DAA treatment eras among individuals living with hepatitis C using a population-based cohort in the province of Ontario, Canada’s most populous jurisdiction and estimated the incidence of liver-death by SVR status for important clinical groups. The availability of real-world population-based data in the current study allows for generalizable findings compared to studies based on hepatology units [22,25]; or those relying on Medicare or U.S. Veteran populations [21,26]. In addition, while many studies focus on all-case death, given the ability to identify condition-specific deaths using linked administrative data, we were able to evaluate the effect of SVR on liver-related deaths [21,22,26]. The current analysis was also able to control for important clinical and demographic confounders such as age, at diagnosis, sex, birth year, socioeconomic factors, viral genotype, liver disease severity, HIV and HBV coinfection, presence of substance use, and other comorbidities. Moreover, we performed stratified analyses to explore moderation effects of liver disease severity and substance-use on SVR and to account for clinical policy changes with respect to the expanded treatment access for these clinical populations between the two treatment eras.

In conclusion, we found achievement of SVR to have a profound impact on reducing liver-related mortality in a population-based sample of individuals living with hepatitis C across a range of liver disease severity. We also found that individuals with a history of drug and/or alcohol related substance use disorder can realise significant benefits in terms of liver-related morality from SVR. However, our findings also show liver disease severity to be a major predictor of liver-related death followed by the presence of substance use disorder, highlighting the importance of early diagnosis and the importance of supporting marginalised individuals in treatment access and supportive programmes such as harm reduction.

## Data Availability

The dataset from this study is held securely in coded form at ICES. While legal data sharing agreements between ICES and data providers (e.g., healthcare organizations and government) prohibit ICES from making the dataset publicly available, access may be granted to those who meet pre-specified criteria for confidential access, available at www.ices.on.ca/DAS. The full dataset creation plan and underlying analytic code are available from the authors upon request, understanding that the computer programs may rely upon coding templates or macros that are unique to ICES and are therefore either inaccessible or may require modification.

## Acknowledgements

Parts or whole of this material are based on data and/or information compiled and provided by Immigration, Refugees and Citizenship Canada (IRCC) current to May 2017. However, the analyses, conclusions, opinions, and statements expressed in the material are those of the author(s), and not necessarily those of IRCC. We thank IQVIA Solutions Canada Inc. for use of their Drug Information File. Parts of this material are based on data and/or information compiled and provided by CIHI. However, the analyses, conclusions, opinions and statements expressed in the material are those of the author(s), and not necessarily those of CIHI. Parts of this report are based on Ontario Registrar General (ORG) information on deaths, the original source of which is ServiceOntario. The views expressed therein are those of the author and do not necessarily reflect those of ORG or the Ministry of Government Services.

## Figure Legends

**Supplemental Table 1.**
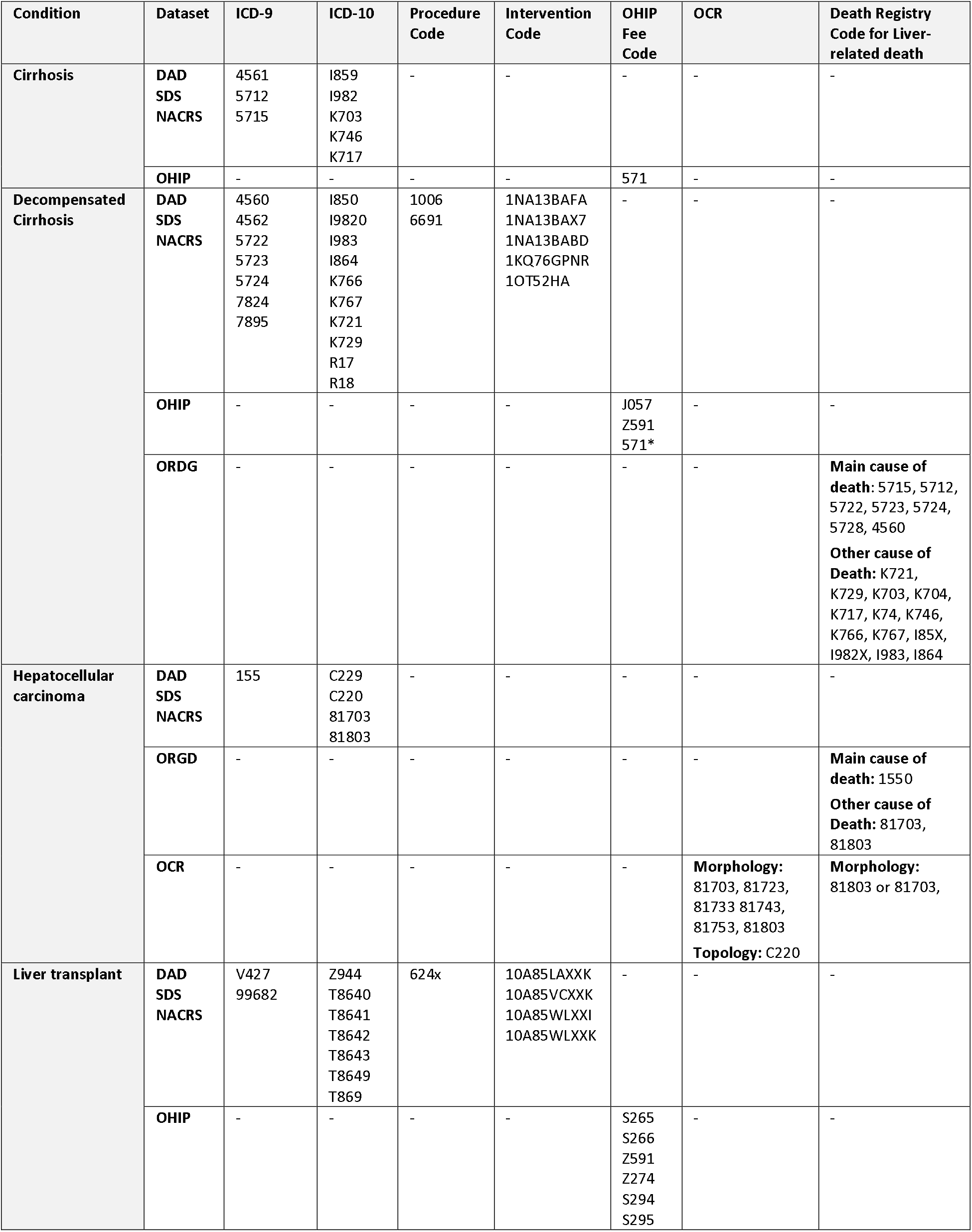
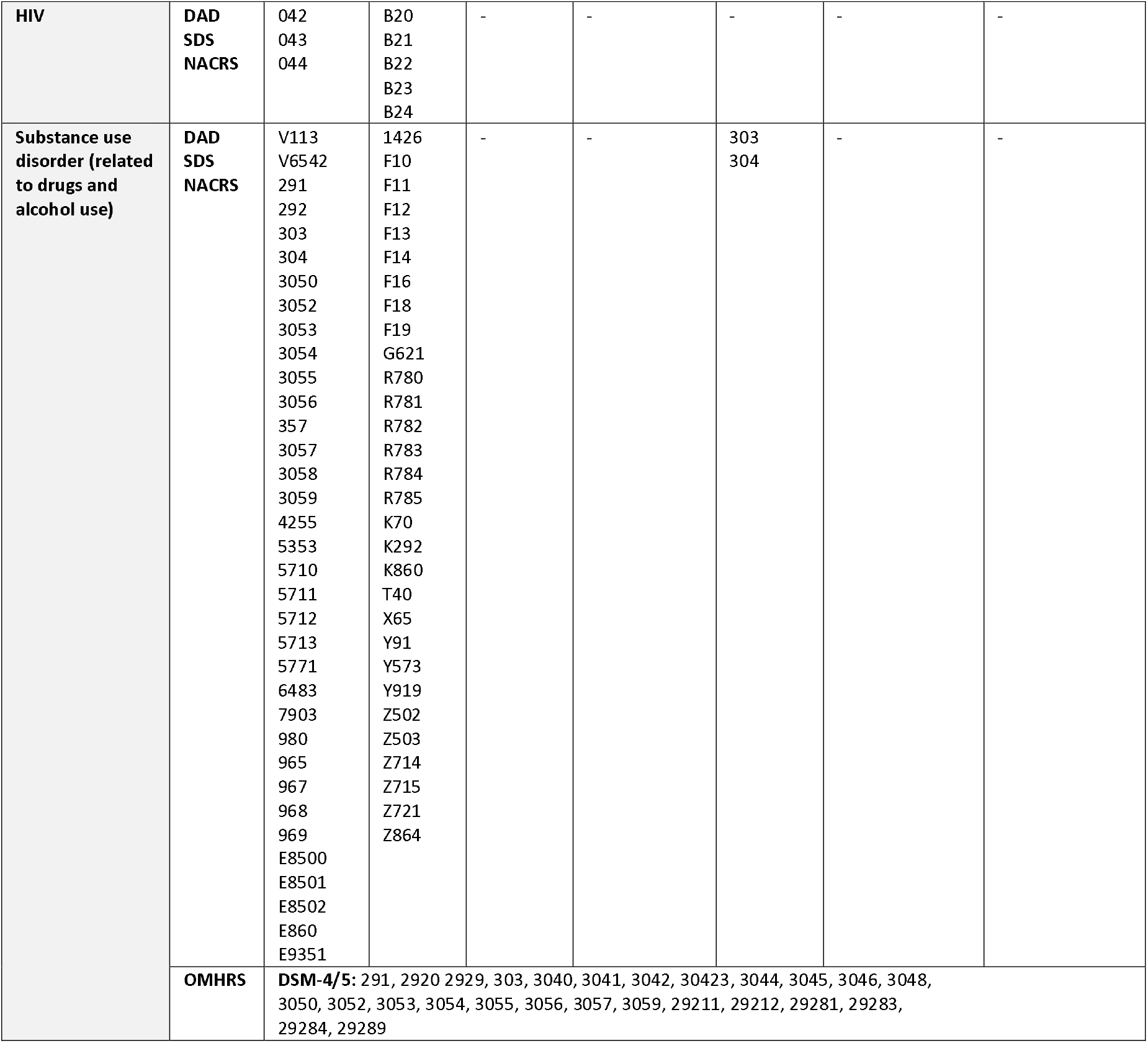
Diagnostic and procedure codes used to identify HCV-related diagnosis and comorbidities from datasets held at ICES. List of diagnostic and procedure codes used to identify HCV-related conditions and comorbidities. Cirrhosis was defined as a single inpatient cirrhosis code listed above. *Decompensated cirrhosis is defined as having cirrhosis using outpatient cirrhosis code (OHIP:571) and at least one inpatient diagnostic code, procedure code or death code associated with decompensated cirrhosis listed above. HCC was defined as either a diagnosis in the Ontario Cancer Registry or inpatient diagnostic code or death code associated hepatocellular carcinoma as listed above. DAD dataset holds diagnostic and procedural data from inpatient hospital admissions. NACRS dataset holds data on diagnostic and procedural information from ambulatory care and emergency department visits. OHIP dataset holds data on claims made by physicians for universally insured services. OCR is the Ontario Cancer Registry and holds information on all Ontario residents with a cancer diagnosis including diagnosis date and details on the type of cancer. The ORGD dataset holds data on the date and cause of death of Ontario residents. The OMHR dataset holds information on individuals receiving adult mental health services in Ontario, and admissions to mental health–designated hospital beds. *Abbreviations: ICD-9: International Classification of Diseases, 9^th^ revision, ICD-10: International Classification of Diseases, 10^th^ revision; NACRS: National Ambulatory Care Reporting System, OHIP: Ontario Health Insurance Program, DAD: Discharge Abstract Database; OCR: Ontario Cancer Registry; ORGD: Office of the Registrar General- Deaths; OMHRS: Ontario Mental Health Reporting System.*

**Supplemental Table 2.**
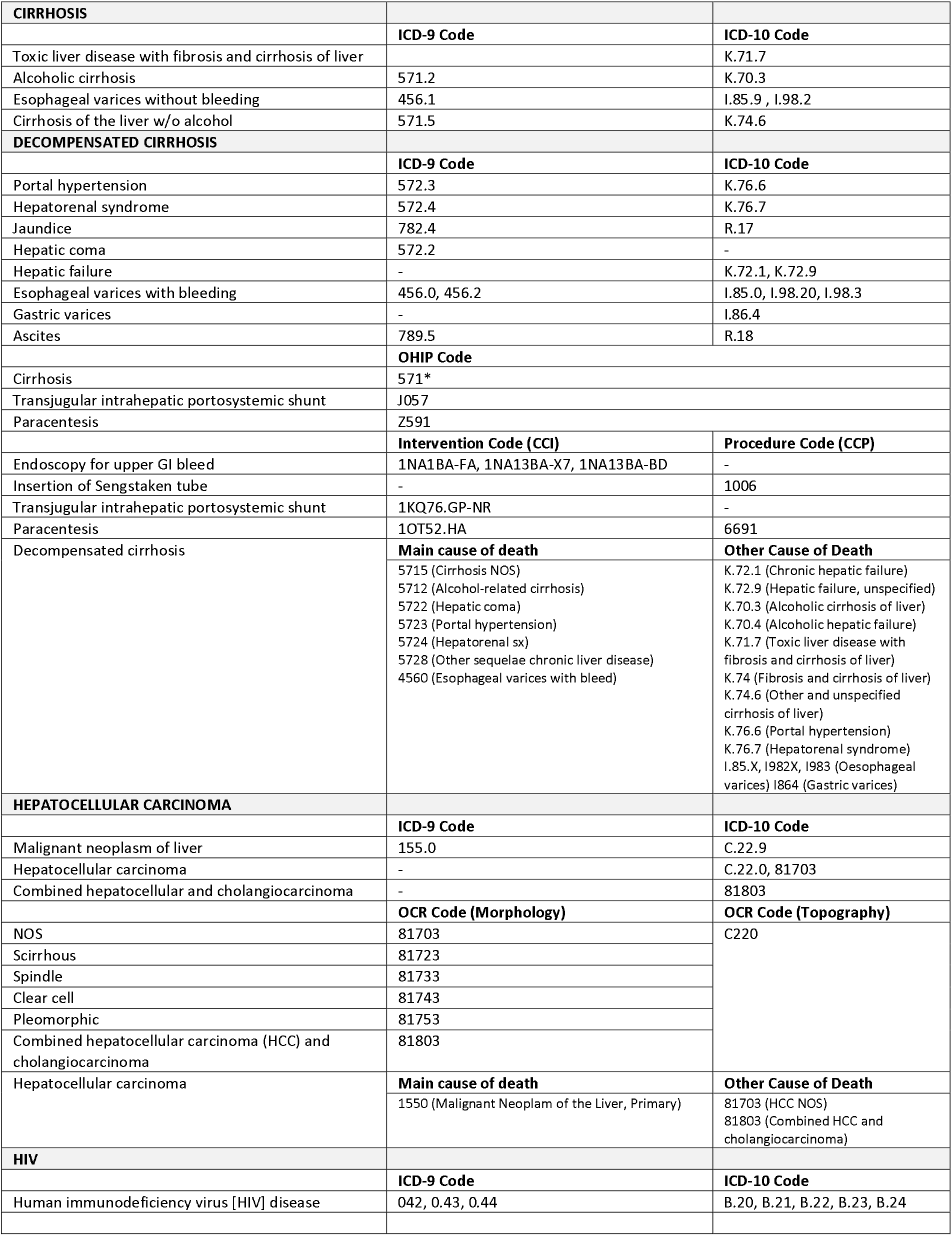
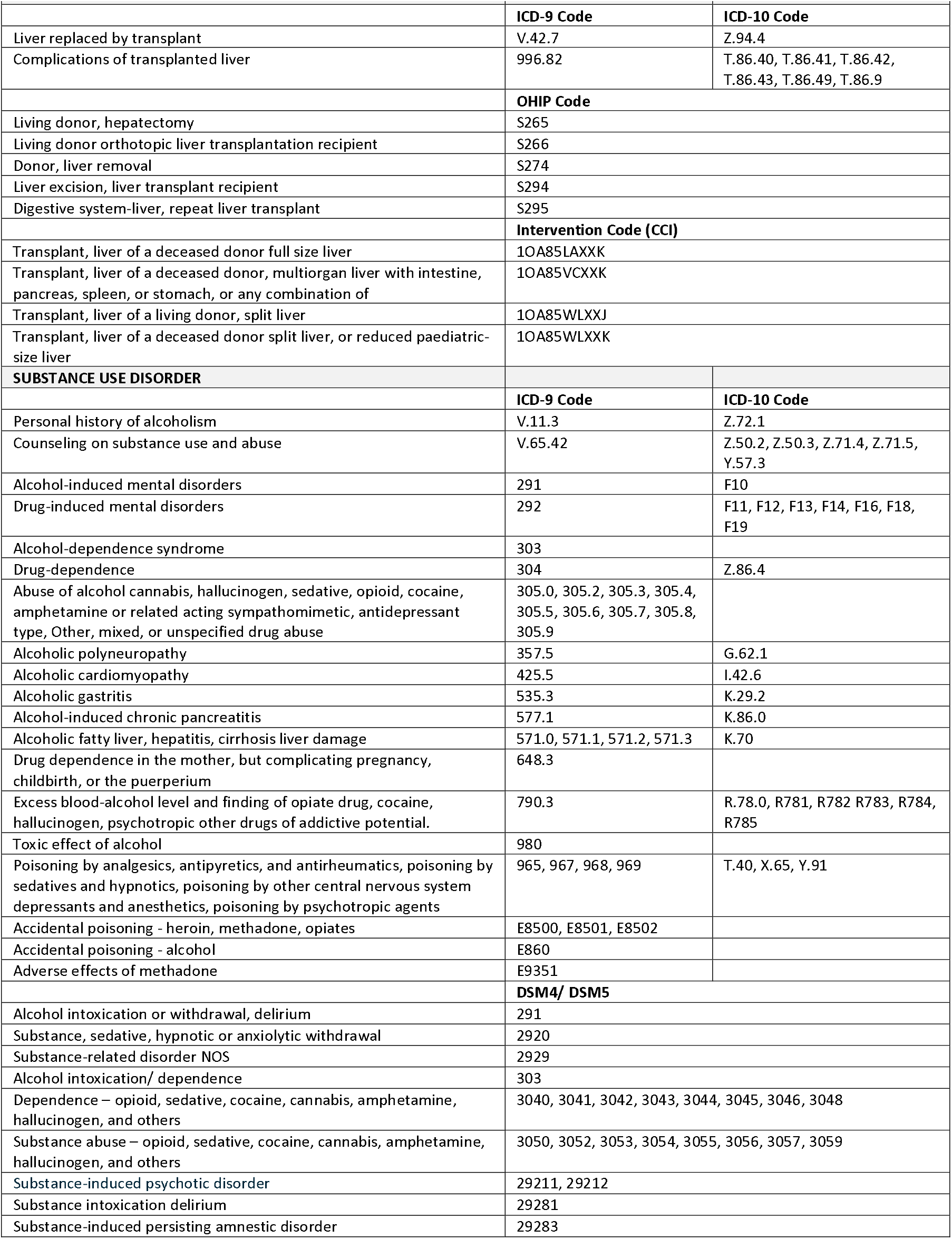
Detailed description of diagnostic, death and procedure-related codes. List of diagnostic and procedure codes used to identify HCV-related conditions and comorbidities. Cirrhosis was defined as a single inpatient cirrhosis code listed above. *Decompensated cirrhosis is defined as having cirrhosis using outpatient cirrhosis code (OHIP:571) and at least one inpatient diagnostic code, procedure code or death code associated with decompensated cirrhosis listed above. *Abbreviations: ICD-9: International Classification of Diseases, 9^th^ revision, ICD- 10: International Classification of Diseases, 10^th^ revision; OHIP: Ontario Health Insurance Program, OCR: Ontario Cancer Registry.*

**Supplemental Table 3.** Drug identification numbers used to identify HCV antiviral treatment. List of DINs used to identify HCV antiviral treatment initiation in the HCV care cascade population. *Abbreviations: DIN: Drug identification number; DAA: direct-acting antivirals; PEG-IFN: pegylated interferon; IFN: Interferon; RBV: ribavirin.*

| DIN | DRUG TYPE | GENERIC NAME (BRAND NAME) |
| --- | --- | --- |
| **DAA-BASED TREATMENTS** |  |  |
| 2370816 | DAA (First generation) | BOCEPREVIR (VICTRELIS) |
| 2371448 | DAA (First generation) | BOCEPREVIR/PEG-INTERFERON ALFA-2B/RIBAVIRIN (VICTRELIS TRIPLE) |
| 2371456 | DAA (First generation) | BOCEPREVIR/PEG-INTERFERON ALFA-2B/RIBAVIRIN (VICTRELIS TRIPLE) |
| 2371464 | DAA (First generation) | BOCEPREVIR/PEG-INTERFERON ALFA-2B/RIBAVIRIN (VICTRELIS TRIPLE) |
| 2371472 | DAA (First generation) | BOCEPREVIR/PEG-INTERFERON ALFA-2B/RIBAVIRIN (VICTRELIS TRIPLE) |
| 2371553 | DAA (First generation) | TELAPRAVIR (INCIVEK) |
| 2416441 | DAA | SIMEPREVIR (GALEXOS) |
| 2418355 | DAA | SOFOSBUVIR (SOVALDI) |
| 2456370 | DAA | SOFOSBUVIR/VELPATASVIR (EPCLUSA) |
| 2467542 | DAA | SOFOSBUVIR/VELPATASVIR/VOXILAPREVIR (VOSEVI) |
| 2432226 | DAA | LEDISPAVIR/SOFOSBUVIR (HARVONI) |
| 2436027 | DAA | DASABUVIR/OMBITASVIR/PARITAPREVIR/RITONAVIR (HOLKIRA PAK) |
| 2447711 | DAA | OMBITASVIR/PARITAPREVIR/RITONAVIR (TECHNIVIE) |
| 2444747 | DAA | DACLATASVIR (DAKLINZA) |
| 2444755 | DAA | DACLATASVIR (DAKLINZA) |
| 2452294 | DAA | ASUNAPREVIR (SUNVEPRA) |
| 2451131 | DAA | ELBASVIR/GRAZOPREVIR (ZEPATIER) |
| 2467550 | DAA | GLECAPREVIR/PIBRENTASVIR (MAVIRET) |
| **RIBAVIRIN** |  |  |
| 2425890 | RBV | RIBAVIRIN (IBAVYR) |
| 2425904 | RBV | RIBAVIRIN (IBAVYR) |
| 2439212 | RBV | RIBAVIRIN (IBAVYR) |
| **INTERFERON-BASED TREATMENTS** |  |  |
| 2242966 | PEG-IFN | PEG-INTERFERON ALFA-2B (UNITRON PEG) |
| 2242967 | PEG-IFN | PEG-INTERFERON ALFA-2B (UNITRON PEG) |
| 2242968 | PEG-IFN | PEG-INTERFERON ALFA-2B (UNITRON PEG) |
| 2242969 | PEG-IFN | PEG-INTERFERON ALFA-2B (UNITRON PEG) |
| 2248077 | PEG-IFN | PEG-INTERFERON ALFA-2A (PEGASYS) |
| 2248078 | PEG-IFN | PEG-INTERFERON ALFA-2A (PEGASYS) |
| 2239730 | IFN/RBV | INTERFERON ALFA-2B/RIBAVIRIN (REBETRON) |
| 2241159 | IFN/RBV | INTERFERON ALFA-2B/RIBAVIRIN (REBETRON) |
| 2253410 | PEG-IFN/RBV | PEG-INTERFERON ALFA-2A/RIBAVIRIN (PEGASYS RBV) |
| 2253429 | PEG-IFN/RBV | PEG-INTERFERON ALFA-2A/RIBAVIRIN (PEGASYS RBV) |
| 2254573 | PEG-IFN/RBV | PEG-INTERFERON ALFA-2B/RIBAVIRIN (PEGETRON REDIPEN) |
| 2246026 | PEG-IFN/RBV | PEG-INTERFERON ALFA-2B/RIBAVIRIN (PEGETRON) |
| 2246027 | PEG-IFN/RBV | PEG-INTERFERON ALFA-2B/RIBAVIRIN (PEGETRON) |
| 2246028 | PEG-IFN/RBV | PEG-INTERFERON ALFA-2B/RIBAVIRIN (PEGETRON) |
| 2246029 | PEG-IFN/RBV | PEG-INTERFERON ALFA-2B/RIBAVIRIN (PEGETRON) |
| 2246030 | PEG-IFN/RBV | PEG-INTERFERON ALFA-2B/RIBAVIRIN (PEGETRON) |
| 2254581 | PEG-IFN/RBV | PEG-INTERFERON ALFA-2B/RIBAVIRIN (PEGETRON) |
| 2254603 | PEG-IFN/RBV | PEG-INTERFERON ALFA-2B/RIBAVIRIN (PEGETRON) |
| 2254638 | PEG-IFN/RBV | PEG-INTERFERON ALFA-2B/RIBAVIRIN (PEGETRON) |
| 2254646 | PEG-IFN/RBV | PEG-INTERFERON ALFA-2B/RIBAVIRIN (PEGETRON) |
| 2223406 | IFN | INTERFERON ALFA-2B (INTRON-A) |
| 2238674 | IFN | INTERFERON ALFA-2B (INTRON-A) |
| 2238675 | IFN | INTERFERON ALFA-2B (INTRON-A) |
| 2240693 | IFN | INTERFERON ALFA-2B (INTRON-A) |
| 2240694 | IFN | INTERFERON ALFA-2B (INTRON-A) |
| 2240695 | IFN | INTERFERON ALFA-2B (INTRON-A) |

**Supplemental Table 4.**
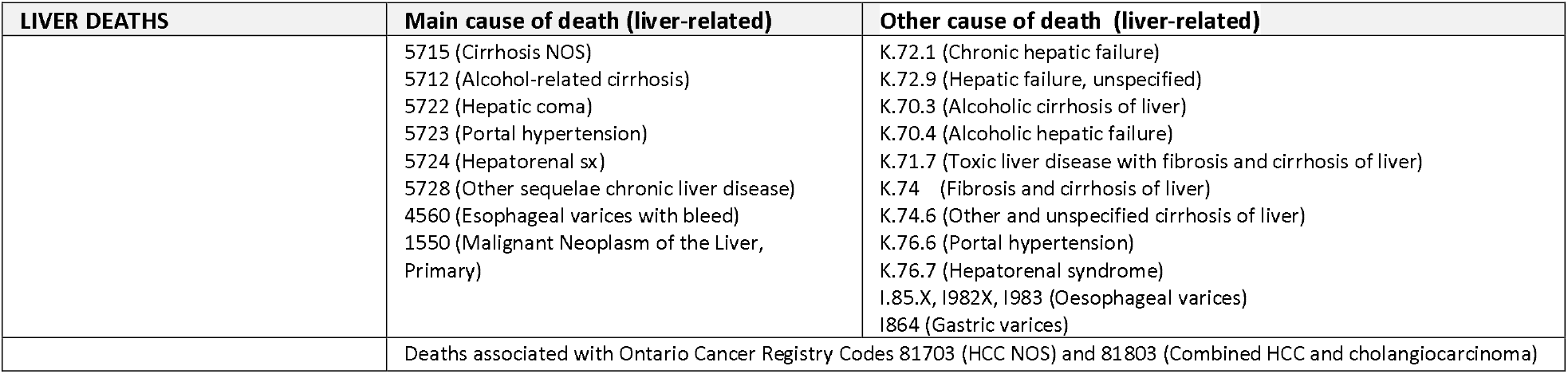
Death code used to identify liver-related death. List of death codes used to identify liver-related deaths. *Abbreviations: HCC: Hepatocellular carcinoma.*

**Supplemental Table 5.**
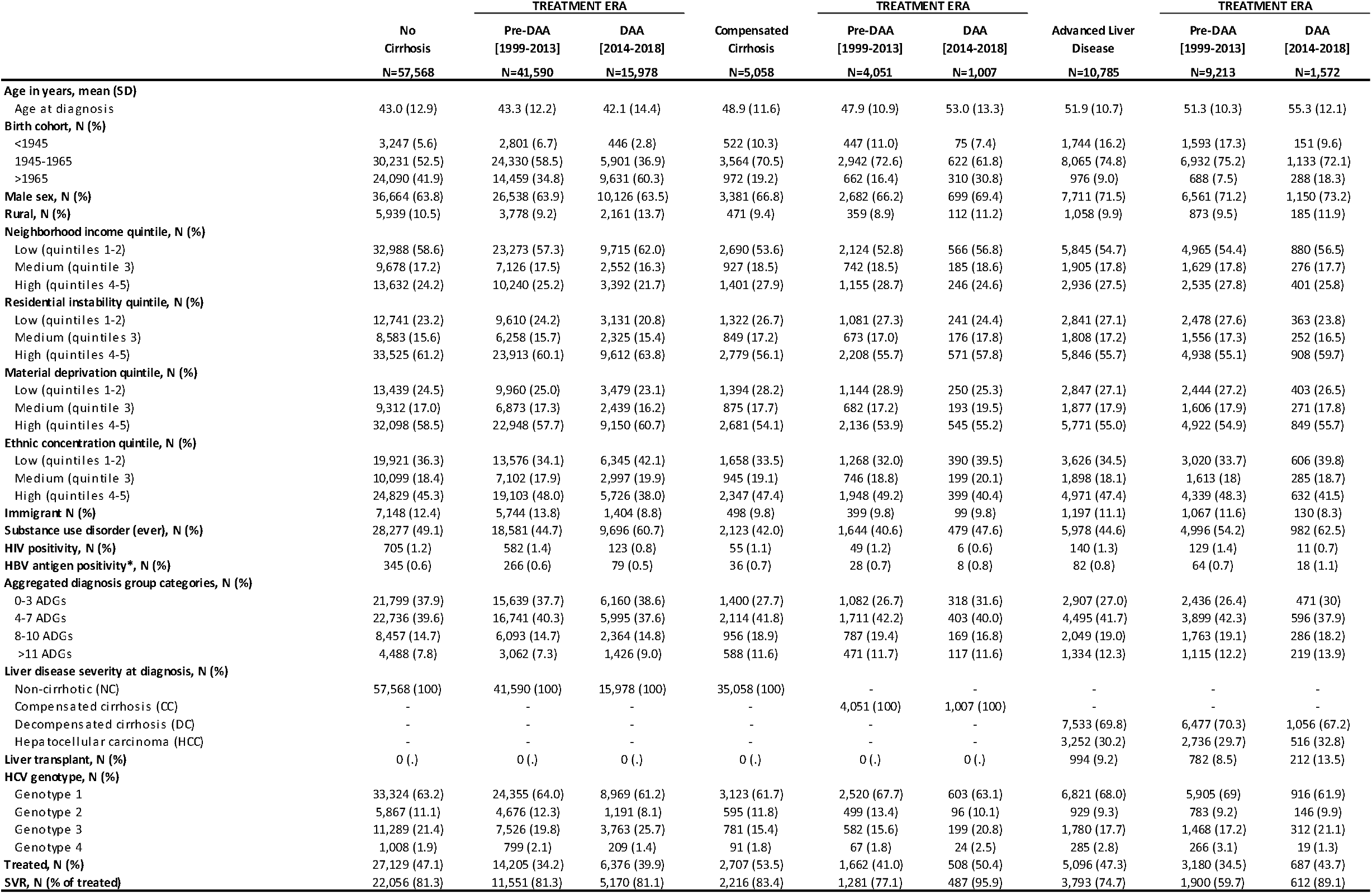
Characteristics of study cohort stratified by liver disease severity and treatment era. Baseline characteristics of the study cohort stratified by liver disease severity at the time of RNA diagnosis for all individuals and for those diagnosed during the pre-DAA era (Jan 1999- Dec 2013) and DAA era (Jan 2014- Dec 2018). Frequencies are calculated after exclusion of missing values. *HBV diagnosis is based on hepatitis b surface antigen (HBsAg) reactivity. *Abbreviations: AB: antibody; RNA: ribonucleic acid; HCV: hepatitis c virus; DAA: direct-acting antiviral; ADG: aggregated diagnostic groups; HBV: hepatitis b virus; NC: no cirrhosis; CC: compensated cirrhosis, DC: decompensated cirrhosis; HCC: hepatocellular carcinoma, HIV: human immunodeficiency virus; HR: hazard ratio; CI: confidence interval; q: quintile; N: number of observations; SVR: sustained viral clearance.*

**Supplemental Table 6.** Characteristics of study cohort stratified by substance use and treatment era. Baseline characteristics of the study cohort stratified by substance use for all individuals and for those diagnosed during the pre-DAA era (Jan 1999- Dec 2013) and DAA era (Jan 2014- Dec 2018). Frequencies are calculated after exclusion of missing values. *HBV diagnosis is based on hepatitis b surface antigen (HBsAg) reactivity. *Abbreviations: AB: antibody; RNA: ribonucleic acid; HCV: hepatitis c virus; DAA: direct-acting antiviral; ADG: aggregated diagnostic groups; HBV: hepatitis b virus; NC: no cirrhosis; CC: compensated cirrhosis, DC: decompensated cirrhosis; HCC: hepatocellular carcinoma, HIV: human immunodeficiency virus; HR: hazard ratio; CI: confidence interval; q: quintile; N: number of observations; SVR: sustained viral clearance.*

|  | No Substance<br>Use Disorder<br>N=37,033 | TREATMENT ERA |  | Substance Use<br>Disorder<br>N=36,378 | TREATMENT ERA |  |
| --- | --- | --- | --- | --- | --- | --- |
|  |  | Pre-DAA<br>[1999-2013]<br>N=29,633 | DAA<br>[2014-2018]<br>N=7,400 |  | Pre-DAA<br>[1999-2013]<br>N=25,221 | DAA<br>[2014-2018]<br>N=11,157 |
| **Age in years, mean (SD)** |  |  |  |  |  |  |
| Age at diagnosis | 48.3 (13.4) | 47.6 (12.9) | 50.9 (15.0) | 41.0 (11.3) | 41.9 (10.6) | 39.1 (12.7) |
| **Birth cohort, N (%)** |  |  |  |  |  |  |
| <1945 | 4,702 (12.7) | 4,115 (13.9) | 587 (7.9) | 811 (2.2) | 726 (2.9) | 85 (0.8) |
| 1945- 1965 | 22,982 (62.1) | 18,796 (63.4) | 4,186 (56.6) | 18,878 (51.9) | 15,408 (61.1) | 3,470 (31.1) |
| >1965 | 9,349 (25.2) | 6,722 (22.7) | 2,627 (35.5) | 16,689 (45.9) | 9,087 (36.0) | 7,602 (68.1) |
| **Male sex, N (%)** | 22,807 (61.7) | 18,111 (61.2) | 4,696 (63.7) | 24,949 (68.6) | 17,670 (70.1) | 7,279 (65.2) |
| **Rural, N (%)** | 3,201 (8.8) | 2,401 (8.3) | 800 (11.0) | 31,795 (88.2) | 2,609 (10.4) | 1,658 (15.0) |
| **Neighborhood income quintile, N (%)** |  |  |  |  |  |  |
| Low (quintiles 1-2) | 18,830 (52.0) | 14,883 (51.4) | 3,947 (54.5) | 22,693 (63.3) | 15,479 (62.3) | 7,214 (65.7) |
| Medium (quintile 3) | 6,851 (18.9) | 5,494 (19.0) | 1,357 (18.7) | 5,659 (15.8) | 4,003 (16.1) | 1,656 (15.1) |
| High (quintiles 4-5) | 10,499 (29.1) | 8,565 (29.6) | 1,934 (26.8) | 7,470 (20.9) | 5,365 (21.6) | 2,105 (19.2) |
| **Residential instability quintile, N (%)** |  |  |  |  |  |  |
| Low (quintiles 1-2) | 10,982 (30.8) | 8,942 (31.3) | 2,040 (28.7) | 5,922 (17.1) | 4,227 (17.5) | 1,695 (16.2) |
| Medium (quintile 3) | 6,071 (17.0) | 4,851 (17.0) | 1,220 (17.1) | 5,169 (14.9) | 3,636 (15.1) | 1,533 (14.7) |
| High (quintiles 4-5) | 18,621 (52.2) | 14,763 (51.7) | 3,858 (54.2) | 23,529 (68.0) | 16,296 (67.4) | 7,233 (69.1) |
| **Material deprivation quintile, N (%)** |  |  |  |  |  |  |
| Low (quintiles 1-2) | 10,545 (29.6) | 8,510 (29.8) | 2,035 (28.6) | 7,135 (20.6) | 5,038 (20.9) | 2,097 (20.0) |
| Medium (quintile 3) | 6,528 (18.3) | 5,243 (18.4) | 1,285 (18.1) | 5,536 (16.0) | 3,918 (16.2) | 1,618 (15.5) |
| High (quintiles 4-5) | 18,601 (52.1) | 14,803 (51.8) | 3,798 (53.3) | 21,949 (63.4) | 15,203 (62.9) | 6,746 (64.5) |
| **Ethnic concentration quintile, N (%)** |  |  |  |  |  |  |
| Low (quintiles 1-2) | 10,854 (30.4) | 8,359 (29.3) | 2,495 (35.1) | 14,351 (41.5) | 9,505 (39.3) | 4,846 (46.3) |
| Medium (quintile 3) | 5,979 (16.8) | 4,703 (16.5) | 1,276 (17.9) | 6,963 (20.1) | 4,758 (19.7) | 2,205 (21.1) |
| High (quintiles 4-5) | 18,841 (52.8) | 15,494 (54.2) | 3,347 (47.0) | 13,306 (38.4) | 9,896 (41.0) | 3,410 (32.6) |
| **Immigrant N (%)** | 7,858 (21.2) | 6,476 (21.9) | 1,382 (18.7) | 985 (2.7) | 734 (2.9) | 251 (2.2) |
| **HIV positivity, N (%)** | 331 (0.9) | 285 (1.0) | 46 (0.6) | 569 (1.6) | 475 (1.9) | 94 (0.8) |
| **HBV antigen positivity\*, N (%)** | 286 (0.8) | 229 (0.8) | 57 (0.8) | 177 (0.5) | 129 (0.5) | 48 (0.4) |
| **Aggregated diagnosis group categories, N (%)** |  |  |  |  |  |  |
| 0-3 ADGs | 15,879 (43.0) | 12,225 (41.3) | 3,654 (49.6) | 10,227 (28.1) | 6,932 (27.5) | 2,679 (28.9) |
| 4-7 ADGs | 14,922 (40.4) | 12,272 (41.5) | 2,650 (36.0) | 14,423 (39.6) | 10,079 (40) | 3,665 (39.6) |
| 8-10 ADGs | 4,435 (12.0) | 3,690 (12.5) | 745 (10.1) | 7,027 (19.3) | 4,953 (19.6) | 1,712 (18.5) |
| >11 ADGs | 1,709 (4.6) | 1,391 (4.7) | 318 (4.3) | 4,701 (13.0) | 3,257 (12.9) | 1,206 (13.0) |
| **Liver disease severity at diagnosis, N (%)** |  |  |  |  |  |  |
| Non-cirrhotic (NC) | 29,291 (79.1) | 23,009 (77.6) | 6,282 (84.9) | 28,277 (77.7) | 20,198 (74.5) | 9,696 (86.9) |
| Compensated cirrhosis (CC) | 2,935 (7.9) | 2,407 (8.1) | 528 (7.1) | 2,123 (5.8) | 1,715 (6.3) | 479 (4.3) |
| Decompensated cirrhosis (DC) | 3,076 (8.3) | 2,745 (9.3) | 331 (4.5) | 4,457 (12.3) | 3,881 (14.3) | 725 (6.5) |
| Hepatocellular carcinoma (HCC) | 1,731 (4.7) | 1,472 (5.0) | 259 (3.5) | 1,521 (4.2) | 1,322 (4.9) | 257 (2.3) |
| **Liver transplant, N (%)** | 438 (1.2) | 361 (1.2) | 77 (1.0) | 556 (1.5) | 447 (1.6) | 135 (1.2) |
| **HCV genotype, N (%)** |  |  |  |  |  |  |
| Genotype 1 | 21,362 (63.0) | 17,176 (63.5) | 4,186 (61.3) | 21,906 (65.3) | 15,604 (67.0) | 6,302 (61.5) |
| Genotype 2 | 4,386 (12.9) | 3,596 (13.3) | 790 (11.6) | 3,005 (9.0) | 2,362 (10.1) | 643 (6.3) |
| Genotype 3 | 5,937 (17.5) | 4,662 (17.2) | 1,275 (18.7) | 7,913 (23.6) | 4,914 (21.1) | 2,999 (29.3) |
| Genotype 4 | 1,221 (3.6) | 1,003 (3.7) | 218 (3.2) | 163 (0.5) | 129 (0.6) | 34 (0.3) |
| Other/mixed | 988 (2.9) | 625 (2.3) | 363 (5.3) | 546 (1.6) | 275 (1.2) | 271 (2.6) |
| **Treated, N (%)** | 19,229 (51.9) | 11,829 (39.9) | 3,478 (47.0) | 15,703 (43.2) | 7,218 (28.6) | 4,093 (35.5) |
| **SVR, N (% of treated)** | 16,629 (86.5) | 9,622 (81.3) | 3,342 (96.1) | 11,481 (73.1) | 5,190 (71.9) | 2,927 (71.5) |

**Supplemental Table 7.** Incidence of clinical events in the study cohort by SVR achievement Liver Disease Severity at Diagnosis Substance Use. Incidence of clinical events per 1,000 person-years in the study cohort for those with and without SVR stratified by liver disease severity at the time of HCV diagnosis and by the presence of absence of substance use disorder. Abbreviations: IR: incidence rate; *SVR: sustained viral clearance*.

|  | Study Cohort | Liver Disease Severity at Diagnosis |  |  | Substance Use |  |
| --- | --- | --- | --- | --- | --- | --- |
|  |  | No Cirrhosis | Compensated Cirrhosis | Advanced Liver Disease | No Substance Use Disorder | Substance Use Disorder |
|  | IR per 1,000 person-years (95%CI) | IR per 1,000 person-years (95%CI) | IR per 1,000 person-years (95%CI) | IR per 1,000 person-years (95%CI) | IR per 1,000 person-years (95%CI) | IR per 1,000 person-years (95%CI) |
| **Liver-related death** | 4.0 (3.8-4.1) | 1.1 (1.0-1.1) | 2.9 (2.5-3.4) | 22.4 (21.5-23.4) | 3.3 (3.1-3.5) | 4.8 (4.5-5.0) |
| No SVR | 5.9 (5.7-6.2) | 1.7 (1.6-1.8) | 5.3 (4.6-6.2) | 34.7 (33.1-36.3) | 5.3 (5.0-5.6) | 6.6 (6.3-7.0) |
| SVR | 1.5 (1.3-1.6) | 0.2 (0.2-0.3) | 0.5 (0.3-0.8) | 9.0 (8.2-9.9) | 1.2 (1.1-1.4) | 1.8 (1.6-2.1) |
| **Non-liver-related death** | 17.4 (17.2-17.7) | 13.9 (13.6-14.2) | 15.5 (14.5-16.5) | 40.3 (39.0-41.6) | 11.6 (11.2-11.9) | 24.8 (24.3-25.3) |
| No SVR | 25.6 (25.1-26.1) | 20.1 (19.7-20.6) | 25.3 (23.6-27.2) | 62.2 (60.1-64.4) | 18.6 (18.0-19.1) | 32.8 (32.1-33.6) |
| SVR | 7.2 (6.9-7.5) | 5.8 (5.5-6.1) | 5.6 (4.8-6.5) | 16.3 (15.2-17.5) | 4.3 (4.1-4.6) | 11.8 (11.3-12.5) |
| **All-cause death** | 21.4 (21.1-21.7) | 15.0 (14.7-15.3) | 18.4 (17.4-19.6) | 62.7 (61.2-64.3) | 14.9 (14.5-15.2) | 29.6 (29.0-30.2) |
| No SVR | 31.5 (31.0-32.4) | 21.8 (21.3-22.3) | 30.6 (28.7-32.7) | 96.9 (94.2-99.6) | 23.9 (23.2-24.5) | 39.4 (35.8-40.2) |
| SVR | 8.7 (8.4-8.9) | 6.0 (5.7-6.3) | 6.0 (5.2-7.0) | 25.4 (24.0-26.8) | 5.6 (5.3-5.9) | 13.7 (13.0-14.3) |

